# A Systems Approach to Assess Transport and Diffusion of Hazardous Airborne Particles in a Large Surgical Suite: Potential Impacts on Viral Airborne Transmission

**DOI:** 10.1101/2020.05.11.20097725

**Authors:** Marc Garbey, Guillaume Joerger, Shannon Furr

## Abstract

Airborne transmission of viruses, such as the coronavirus 2 (SARS-CoV-2), in hospital systems are under debate: it has been shown that transmission of SARS-CoV-2 virus goes beyond droplet dynamics that is limited to 3-6 feet, but it is unclear if the airborne viral load is significant enough to ensure transmission of the disease. Surgical smoke can act as a carrier for tissue particles, viruses, and bacteria. To quantify airborne transmission from a physical point of view, we consider surgical smoke produced by thermal destruction of tissue during the use of electrosurgical instruments as a marker of airborne particle diffusion-transportation. Surgical smoke plumes are also known to be dangerous for human health, especially to surgical staff who receive long-term exposure over the years. There are limited quantified metrics reported on long-term effects of surgical smoke on staff’s health. The purpose of this paper is to provide a mathematical framework and experimental protocol to assess the transport and diffusion of hazardous airborne particles in every large operating room suite. Measurements from a network of air quality sensors gathered during a clinical study provide validation for the main part of the model. Overall, the model estimates staff exposure to airborne contamination from surgical smoke and biological material. To address the clinical implication over a long period of time, the systems approach is built upon previous work on multi-scale modeling of surgical flow in a large operating room suite and takes into account human behavior factors.

## Introduction

There is a large debate on the possible airborne transmission of coronavirus 2 (SARS-CoV-2) in closed buildings [1,2]. In the unfortunate case an elective surgery is practiced on an asymptomatic COVID-19 patient and who was not tested positive, one may ask if the virus can escape an Operating Room (OR) kept under positive pressure and expose staff in peripheral area to the disease. This question is particularly important to healthcare staff who spend multiple long hour shifts in a hospital system that manages COVID-19 patients. To quantify airborne transmission from a *physical point of view*, we consider surgical smoke as a marker of airborne particle diffusion-transportation emitted from the surgical table area.

Surgical smoke is 95% water or steam and 5% particle material and therefore surgical smoke can act as a carrier for tissue particles, viruses, and bacteria [3]. Today, the risk of surgical smoke has clearly been established [4–10]. One of the main difficulties is that surgical smoke carries Ultra Fine Particles (UFP) as small as 0.01 *microns*, which are able to bypass pulmonary filtration, and small particles up to several *microns* [9]. It was recently shown in a study that air quality, especially concentration of fine particles, is associated with an increase in COVID-19 mortality [11]. Respiratory protection devices are used to protect staff in healthcare facilities with various degrees of success [12,13].

We propose to construct a rigorous multi-scale computational framework to address these questions and use measurements of diffusion-transportation of surgical smoke particles with off-the-shelf portable sensors to calibrate the model.

This methodology addresses only the physical side of the problem and therefore does not answer the effectiveness of airborne particles to induce COVID-19. Some of the difficulties encountered in such studies are that air sampling and infection may or may not be strongly correlated [14,15]. However, it is an important step to quantify the level of exposure in order to estimate the corresponding viral load in part. Transport and diffusion mechanisms are very effective for UFP to travel a long distance from the source in a short period of time. A 2020 report from China demonstrated that SARS-CoV-2 virus particles could be found in the ventilation systems in restaurants [16] and in hospital rooms of patients with COVID-19 underlining how viable virus particles can travel long distances from patients [17].

Clinical environments are too complex to model with the traditional modeling method of airflow and particle transportation because both the source intensity of surgical smoke [18] as well as the mechanism of propagation via door openings [19] are largely dominated by human factors. The geometric complexity of the infrastructure and of the heating, ventilation, and air conditioning (HVAC) system limit the capability of Computational Fluid Dynamics (CFD) [19,19–25] to predict indoor air quality and health [26]. Last but not least, droplet behavior depends not only on their size, but also on the degree of turbulence and speed of the gas cloud, coupled with the properties of the ambient environment (temperature, humidity, and airflow) [2].

We present in this paper a mathematical framework and experimental protocol to assess the transport and diffusion of hazardous airborne particles in any large OR suite. Human behavior factors are taken into account by using a systems and cyber-infrastructure approach [27–29] coupled to a multi-scale modeling of surgical flow in a large OR suite [30]. Overall, the model estimates staff’s exposure to airborne contamination, such as surgical smoke or biological hazard. Validation is provided by a network of wireless air quality sensors placed at critical locations in an OR suite during the initial phase of the surgical-suite-specific study.

A step-by-step construction of the model scaling up from the OR scale to the surgical suite scale will be presented; the model integrates the transport mechanism occurring at the minute scale with the surgical workflow efficiency simulation over a one year period. To assess potential contamination from one OR to another, the extent of the propagation of surgical smoke in the area adjacent to the OR will be checked – this might be more significant than the level of concentration itself.

## Results

Following are the results on the circulation of surgical smoke in a surgical suite starting from a local source of emission in the OR and ending on global dispersion in the suite. A detailed CFD model of the airflow in the OR along with its immediate adjacent structure will be used to build an upper-scale, simplified model. A series of air quality measurements based on the density of particles at specific locations will be used for calibration of the model and for validation purposes.

### Measuring Sources of Surgical Smoke

The measured rate of particles generated by various energy sources, such as monopolar cautery, argon plasma coagulation (APC), and harmonic sources, are found by testing them in an OR space allocated to training, i.e. without patients. The unit used for the source of the emission is 0.01 particles per cubic foot; it gives the measurements an order of magnitude from ten to thousands by which they can be compared. Small particles are found in the range of 0.5 to 2.5 *microns*, which are the sizes of biological-material particles like viruses. As opposed to the results reported in Weld et Al [31], our off-the-shelf particle sensor does not give us access to the UFP count. A conservative estimate from Weld et Al [31] results would be that the concentration number of UFP is 2 to 3 orders of magnitude larger than what is measured for the small particles. In Table 1, each source’s mean, standard deviation, and diffusion coefficient are reported – more precisely *α* is the rate at which the pollutant concentration decreases, which is obtained from fitting a simple exponential decay model *Source* exp^−^*^αt^* to the experimental data.

**Table 1.** Comparison of amplitudes and diffusion coefficients for commonly used electrosurgical instruments.

| **Amplitude (.01 PpCF)** | **Monopolar** | **APC** | **Harmonic** |
| --- | --- | --- | --- |
| **Mean** | 4800 | 1600 | 1050 |
| **Standard Deviation** | 5100 | 1100 | 870 |
| **Diffusion Coefficient** | **Monopolar** | **APC** | **Harmonic** |
| **Mean** | 0.65 | 0.56 | 0.36 |
| **Standard Deviation** | 0.13 | 0.19 | 0.3 |
| **Number of Experiments** | 5 | 5 | 4 |

There was no significant statistical difference between the rates of diffusion of the particles emitted by the monopolar versus the APC instruments. The coefficient of diffusion corresponding to the harmonic instrument is lower but has strong variation. We interpret this result with the fact that some of the particles emitted in this case were too small, 0.06 *microns*, to be detected by our sensors [31]. As mentioned earlier, covering the sensor with a surgical facemask dropped the number of large particles to some extent, but there are always leaks on the sides. As noticed in the literature standard, surgical facemasks do not protect from UFP.

### Airborne Transport-Diffusion at the Operating Room Scale

A 3-Dimensional (3D) CFD model is used to simulate the dispersion of a single source of pollutant in an OR. The dimensions used in the model are the ones in the surgical suite where the clinical study and validations were made. Every OR is different, but the order of magnitude of the physical quantities is the same for each OR in our clinical study. Figure 1 provides the geometric details of the simulation setup that takes into account the geometry of the room, location of air conditioning ducts, location of the doors, and air leaks due to positive pressure despite closed doors.

**Figure 1.**
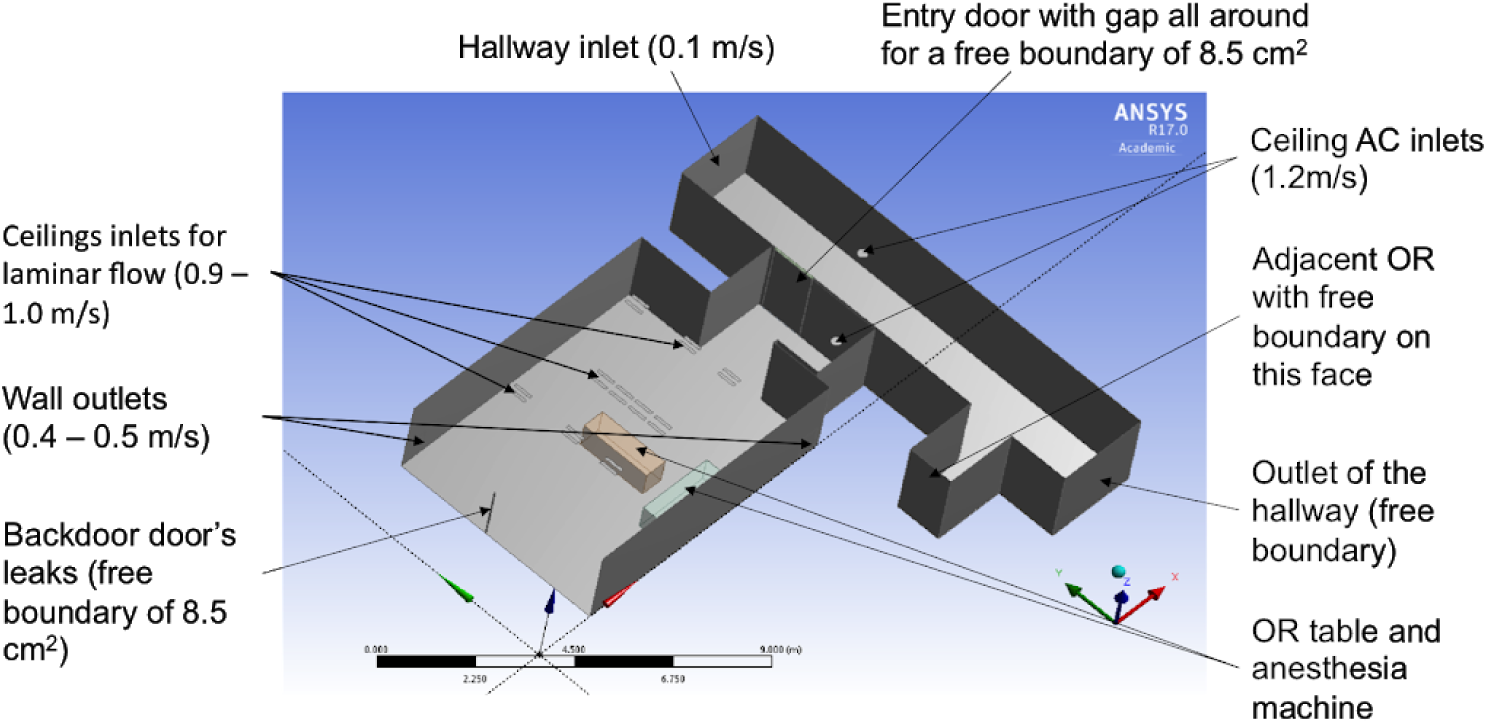
3-Dimensional model of the OR and adjacent hallway.

Table 2 lists the boundary conditions on velocities and temperatures of the OR and its adjacent hallway obtained from measurements.

**Table 2.**
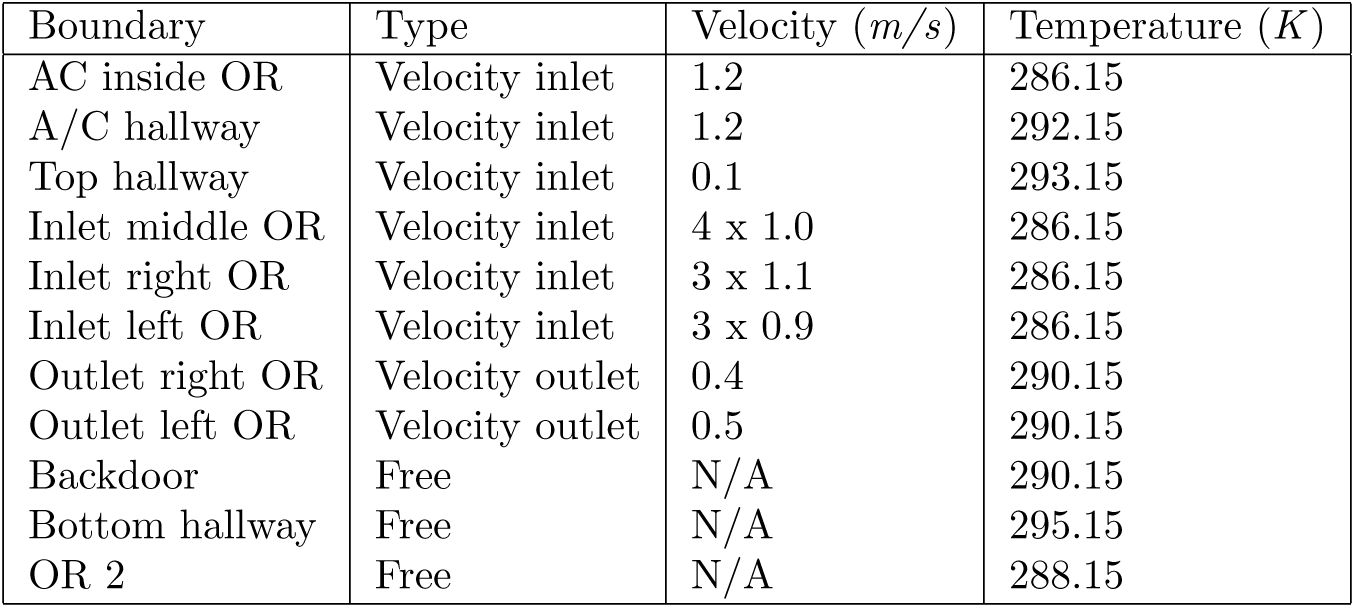
Boundaries conditions of our system with velocities and temperatures.

| Boundary | Type | Velocity ($m/s$) | Temperature ($K$) |
| --- | --- | --- | --- |
| AC inside OR | Velocity inlet | 1.2 | 286.15 |
| A/C hallway | Velocity inlet | 1.2 | 292.15 |
| Top hallway | Velocity inlet | 0.1 | 293.15 |
| Inlet middle OR | Velocity inlet | 4 x 1.0 | 286.15 |
| Inlet right OR | Velocity inlet | 3 x 1.1 | 286.15 |
| Inlet left OR | Velocity inlet | 3 x 0.9 | 286.15 |
| Outlet right OR | Velocity outlet | 0.4 | 290.15 |
| Outlet left OR | Velocity outlet | 0.5 | 290.15 |
| Backdoor | Free | N/A | 290.15 |
| Bottom hallway | Free | N/A | 295.15 |
| OR 2 | Free | N/A | 288.15 |

The surgical smoke plume in the CFD model was simulated using an injection of CO_2_ at the location of the OR table for a duration of 10 seconds. The CO_2_ phase was tracked in the multi-phase CFD simulation as a marker of pollution. The smaller the particle, the best the dispersion model would be based on gas transportation. Figure 2 shows the dispersion of the plume inside and outside the OR, while the door is closed. Dispersion into the hallway was due to the air leaks between the door of the OR and its frame. Verification of the simulation was obtained by refining the mesh and time step until it reached a numerical convergence on the quantities of interest – in particular, the density of CO_2_ and velocity of flow at specific locations.

**Figure 2.**
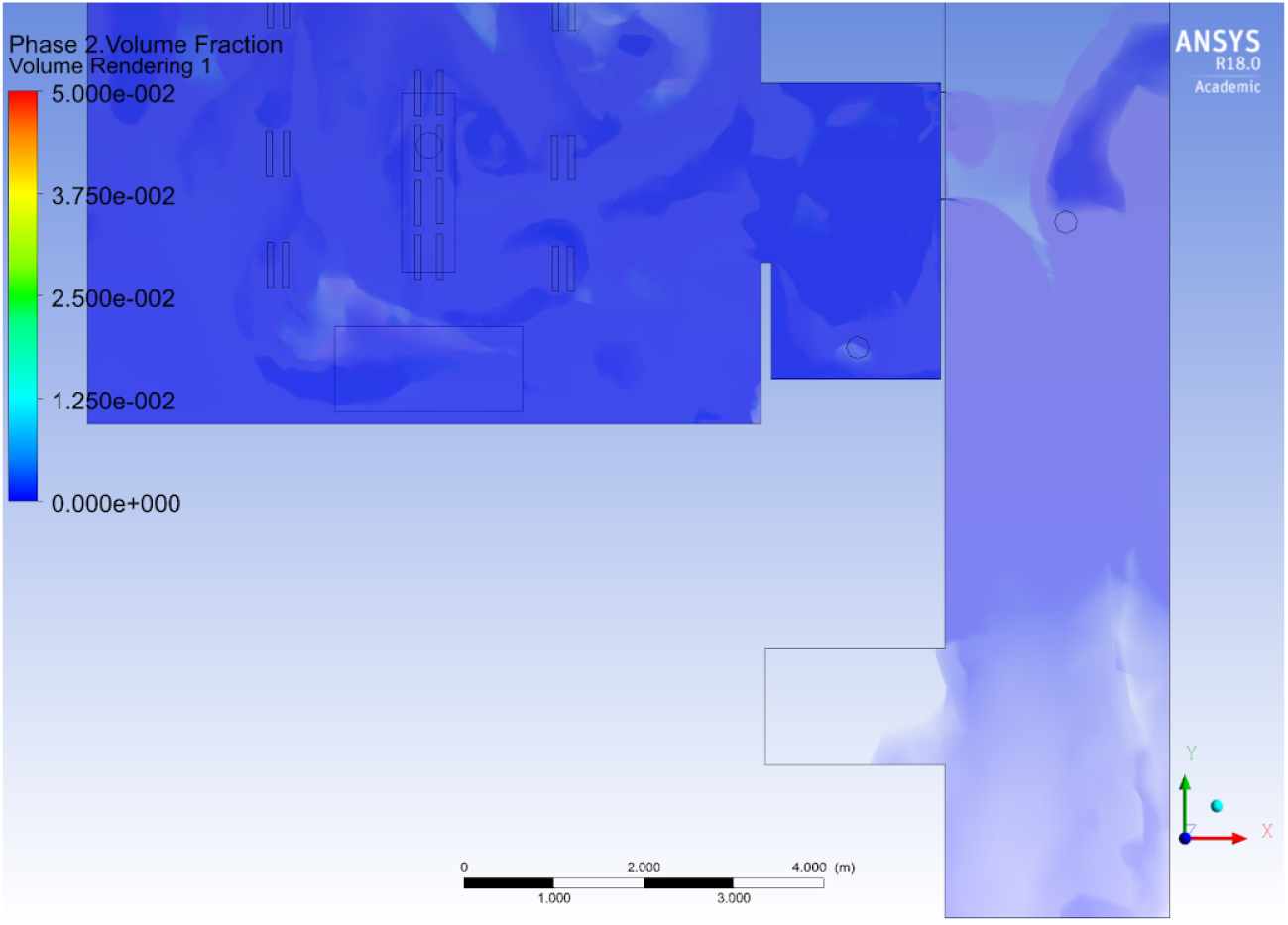
CFD simulation tracking the CO_2_ source emitted at the OR table; in the CFD simulation, injection is done for 10 seconds at the beginning of the simulation. The door of the OR is closed, while the door of the adjacent OR is open. This figure shows that the plume reaches the adjacent OR after 130 sec.

Table 3 provides a comparison between the different velocity values found by the model and by the direct measurement obtained at those locations. The time intervals were also computed: between the emission of the pollutant and the time when an air sensor detected the pollutant inside the OR, close to the door, and in the hall outside the door. The results of Table 3 provides the first level of validation of the CFD simulation.

**Table 3.** Comparison of different values found by the model simulation and by measurement during experimental tests.

| Parameter | Measurement | Simulation |
| --- | --- | --- |
| Velocities (inside leaks in $m/s$) | 3.55 [32] | 3.6 |
| Velocities (in front of the leaks in $m/s$) | 1.5 | 1.2 |
| Velocities (in front of the door in $m/s$) | 0.8 | 0.8 |
| Pressure ($Pa$) | 7.5 | 7.3 |
| $R_1^3$ | 0.56 | 0.6 |
| Delay sensor 3 ($sec$) | 10.33 | 10 |
| Delay sensor 1 ($sec$) | 31 | 29 |

In Table 3, 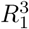 is the ratio of pollutant phase concentration between the sensor location 1 and 3 in the OR in Figure 3, and is interpreted as a small particle density ratio as well.

**Figure 3.**
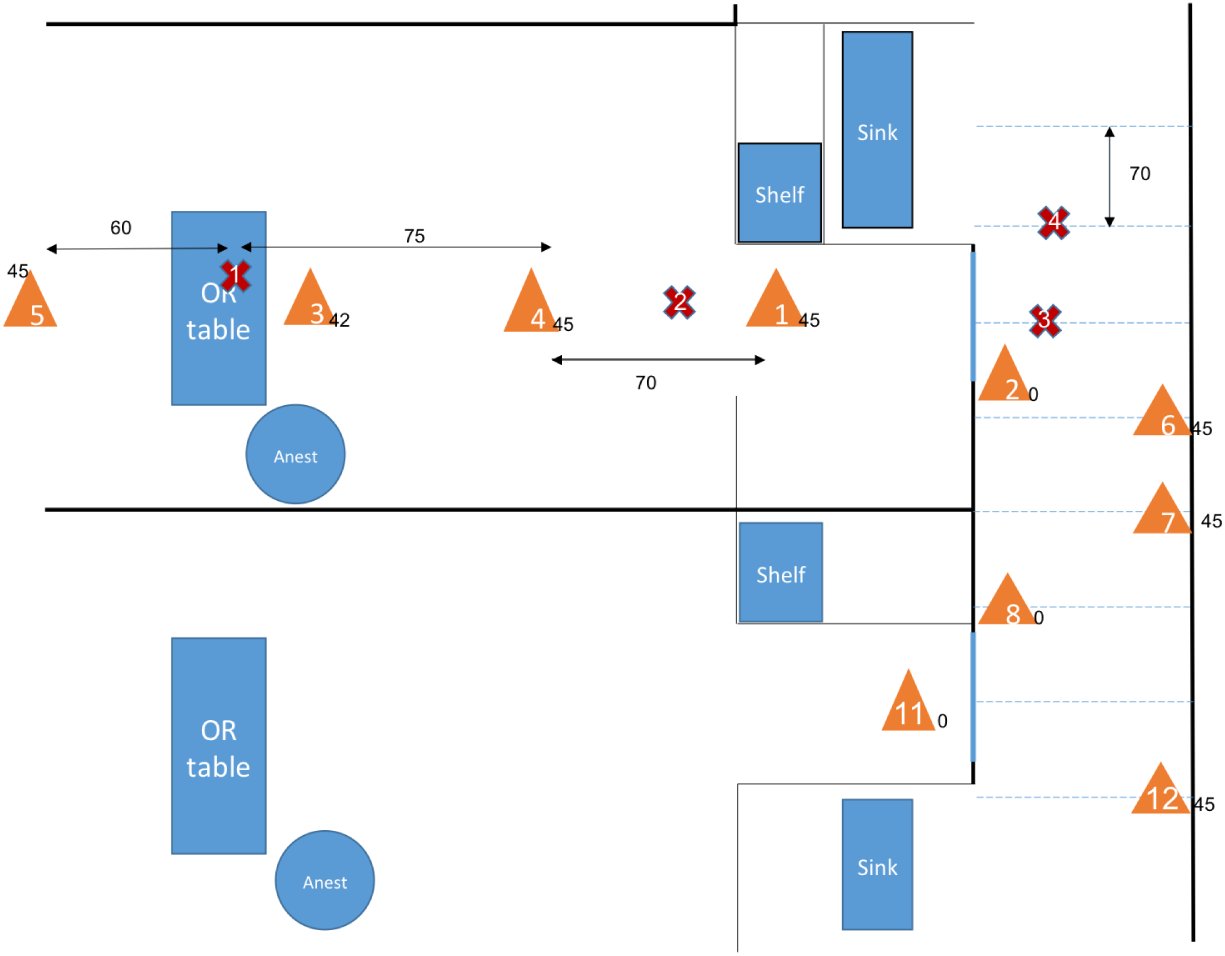
Position of the sensors and sources during the experiments with different source locations. Only one source location was used for each experiment. Sensors are represented by orange triangles and sources by red crosses. The numbers next to the triangles are the heights of the sensors.

**Figure 4.**
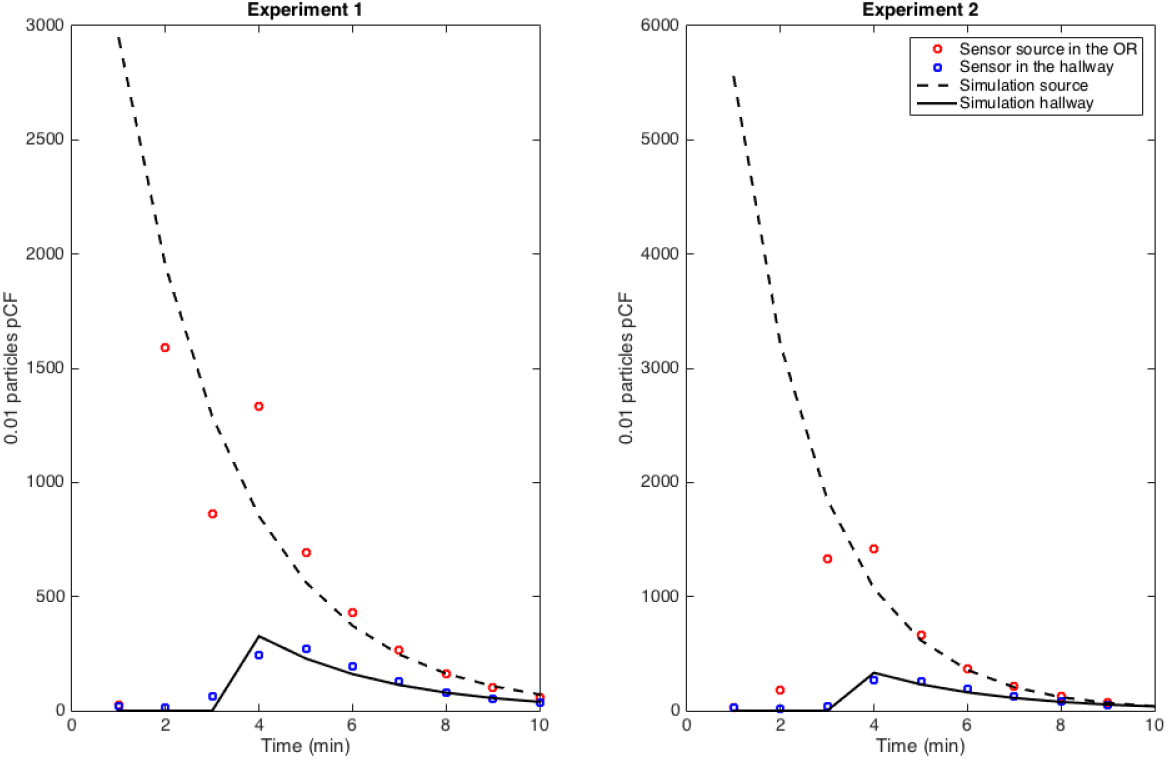
Source in closed-door OR and its impact on hallway air concentration: a 2 *min* delay in transmission from OR to hallway and an exponential decay for each signal was observed.

**Figure 5.**
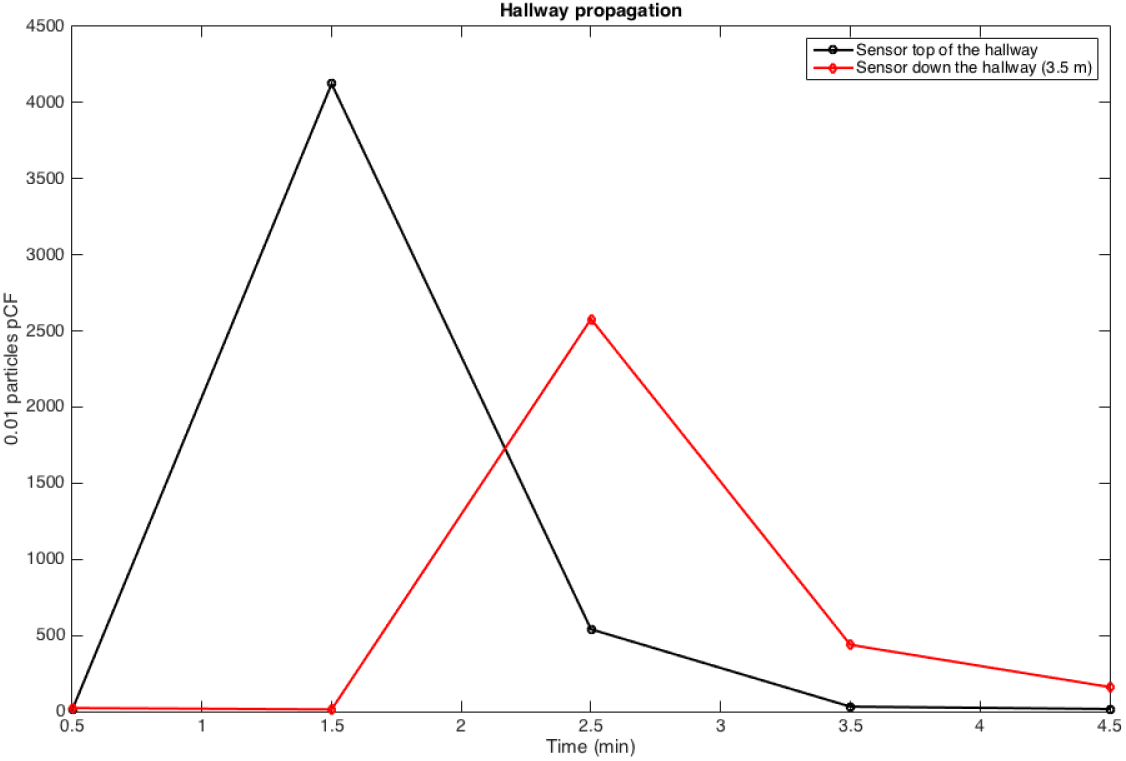
Source in closed-hallway and its propagation down the hallway at marker 6 and 12 locations in Figure 3: convection of the signal down the hallway and an exponential dampening of the signal was observed.

It is particularly interesting to notice that the flow at the door has a 3-dimensional component that is driven by the pressure gradient as well as the temperature difference between the OR and the hallway. While the OR is kept under positive pressure, it loses this pressure as soon as the door is opened. Because the temperature of the OR is generally cooler than the temperature of the hall, we observe from the CFD that the buoyancy effect causes back-flow between the adjacent hallway when the door is opened. This might be part of the mechanism of contamination between ORs. This result is consistent with air quality measurements done in controlled experimental conditions presented hereafter.

It was found that the mixing of contaminants from a burst source to the rest of the OR is reached within a minute; applying a simplified compartment model to describe the OR’s contribution to pollutants using a time step of one minute became apparent. This upper-scale model described in the methodology section will be calibrated next.

### Transmission Condition and Transport-Diffusion of Particles at the Surgical Suite scale

The identification of the model parameters from the experimental data-set corresponding to the setup in Figure 3 is explained below. The experiment was designed first, to assess the delay of pollutant transmission between the OR and the hallway depending on if the door was opened or closed. Second, to compute the rate at which pollutant concentration decreases. An exponential decay was observed in the OR, which is consistent with the fact that diffusion is the main mechanism. However, the hallway behaves more like a duct with a combination of convection and diffusion running down the hallway. These measurements are consistent with the CFD simulation results shown previously. Fitting the simplified model to the controlled experiment with a single source of smoke, the coefficients of diffusion in the OR and in the hallway can be retrieved, as well as the convection velocity in the hallway – see Table 4.

**Table 4.**
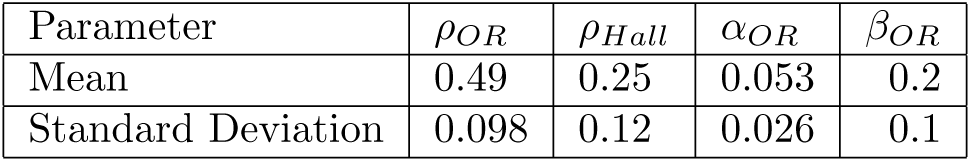
Parameters of the model obtained by fitting the outcome on single-source controlled experiments with injection source locations; 9 measures were used to obtain this table. The coefficients of decay are (*ρ*_OR_ and *ρ_Hall_*) as well as the coefficients of transmission between these spaces (*α_OR_*) from the OR to the hall. *β_OR_* represents the flow from the OR to the hall when the door is open.

| Parameter | $\rho_{OR}$ | $\rho_{Hall}$ | $\alpha_{OR}$ | $\beta_{OR}$ |
| --- | --- | --- | --- | --- |
| Mean | 0.49 | 0.25 | 0.053 | 0.2 |
| Standard Deviation | 0.098 | 0.12 | 0.026 | 0.1 |

The diffusion coefficient in the OR and the hallway are dependent on the HVAC system that is, by design, more effective in the OR than in the hallway. Therefore, the rate of decay in the OR is twice as large as the rate of the decay in the hallway. As reported before, the diffusion coefficient for the particle tracking setting is about the same for the spray source as it is for the monopolar or APC sources. The transmission condition with a closed OR door is not negligible: it is about 4 times less than with an open door.

From Figure 1, the traveling wave velocity is reconstructed and travels about one OR width in a minute, *v*_0_ is about 0.1 *m/s* at mid-hall location. This small velocity in the hallway could not be directly measured, but it is in agreement with the CFD simulation reported earlier.

The most important result is summarized in Figure 6: surgical smoke emitted in a single OR can rapidly reach the hallway within a minute due to the OR door opening and is diluted by a factor of roughly 10. In the unfortunate event that the door of the next OR is opened, then some trace of the surgical smoke emitted by the OR upstream, can flow *inside the next OR down the hall*; while the level of exposure to surgical smoke would be insignificant in this second OR, it is clear that the standard positive pressure established in these ORs cannot guarantee those airborne particles do not propagate from one OR to another. Over a period of several months, this rare event might be capable of propagating an airborne disease. In fact, the frequency of door openings of each OR can be very high, as shown in Figure 14, that the probability of propagating airborne diseases and contaminating other ORs seems inevitable. Next, to systematically assess long-term exposure, the result obtained by coupling the air quality model with an Agent-Based Model (ABM) of the surgical flow will be reported on.

**Figure 6.**
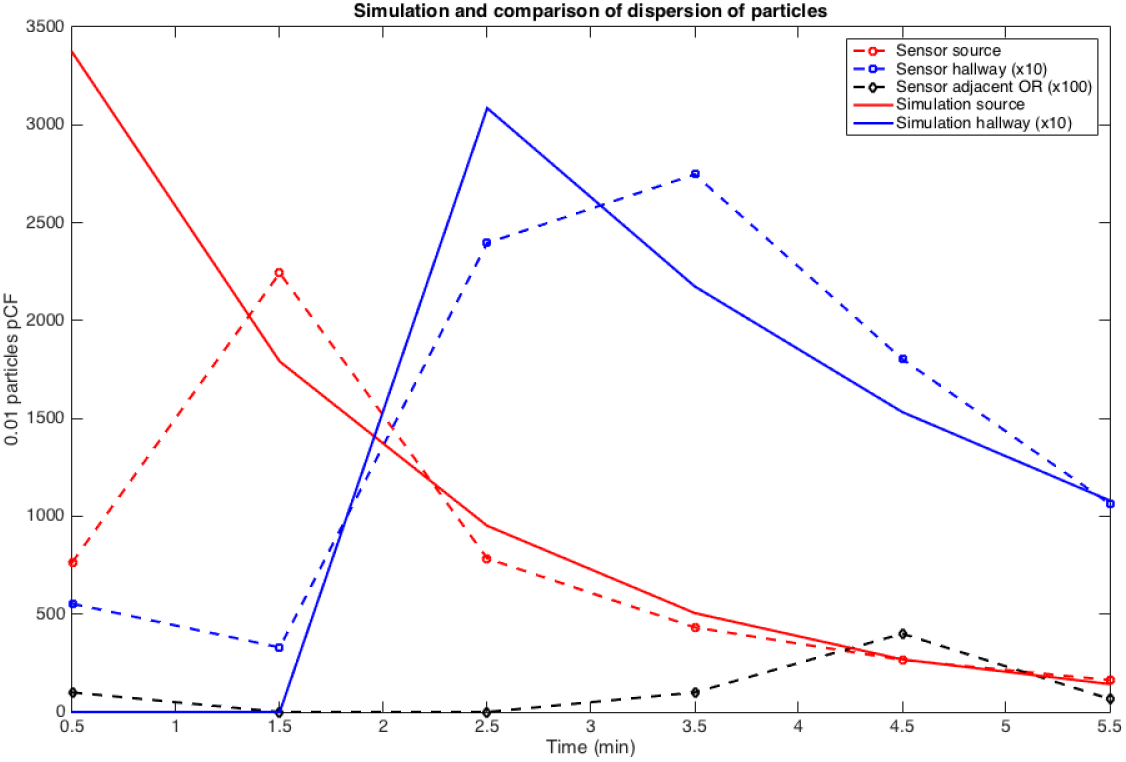
Effect of door opening and closing on propagation of marker from one OR to the next OR down the hall: ○ is the sensor close to the source in the OR, □ is 10 times the concentration down the hall, and ⋄ is 100 times the measured concentration in the next OR down the hall – for convenience, we have plotted in solid line the exponential model fitting for the experimental datasets in the main OR and in the hall.

### Clinical Agent-Based Model of Hazardous Airborne Particles

Figure 7 shows a measurement done during a clinical study with 3 consecutive laparoscopic procedures during the day. The red curve accounts for the number of particles detected by the sensor inside the OR, while the blue curve provides the corresponding measurement from the hallway. Patients’ registration starts at 7 a.m., before any surgery occurs, and lasts all day until all surgeries are complete. Large peaks of particle concentrations were observed during the times the OR was being cleaned. These peaks were removed from the OR acquisition curve that corresponds to the use of detergent. Similarly, during the preparation and closing of the patient, the sensor sometimes captured the use of chemicals when preparing the sterile field or the leak of anesthetic gas. The red peak during the third procedure in Figure 7 most likely corresponds to an excess of surgical smoke. As expected, the concentration of particles in the hall is not strictly correlated to the emission of surgical smoke in the OR. The hallway collects pollutants from a number of ORs under positive pressure at the same time. Because of this, it is difficult to separate out surgical smoke from other sources in the hallway measurement, such as chemicals used in the preparation of patients located in ORs upstream. The model is built to qualitatively reproduce the concentration of surgical smoke inside an OR and its adjacent section of the hall.

**Figure 7.**
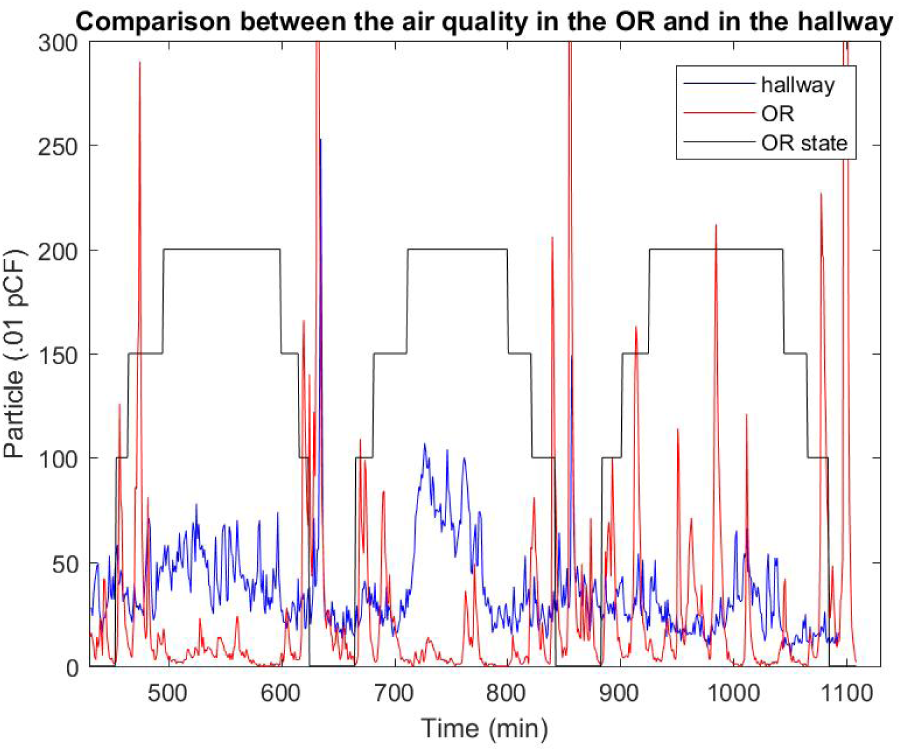
Measurement of the air quality in the OR and in the hallway compared with the status of the surgery. Here, three laparoscopic surgeries are analysed. The steps follow: Patient in (100) – Patient extubated (150) – Start laparoscopic portion (200) – Stop laparoscopic portio – Patient extubated – Patient out.

In our simulation (see Figure 8), the emission of surgical smoke is restricted during the time the patient is intubated. This simulation was done on the whole surgical suite with a stochastic production of smoke in each OR similar to the one reported in Meeusen et al. [18]. One observation is the same pattern of pollutant concentration in the hallway as seen in the clinical dataset. In particular, there is no obvious correlation between the source of smoke in the OR of that simulation and the concentration in the section of the adjacent hallway. In fact, while exposure to surgical smoke in the OR is relatively intense in a short period of time and then vanishes, the pollutant is stagnant in the hallway for a much longer period of time and therefore contributes to long-term exposure. Overall, the delay Δ*t*_2_ in the transmission conditions in (4) and (5) has very little influence on the result and can be neglected.

**Figure 8.**
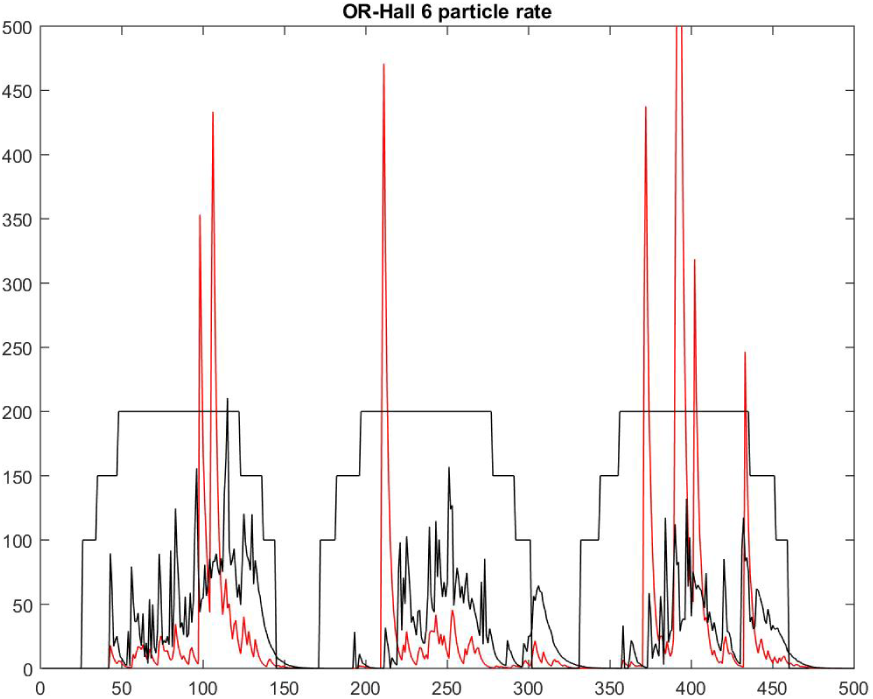
Result of the simulation of three surgeries using the model with stochastic production of smoke and opening of door linked to the status of the surgery. The steps follow: Patient in (100) – Patient extubated (150) – Start laparoscopic portion (200) – Stop laparoscopic portio – Patient extubated – Patient out.

To expand the study, this simplified air quality model was then coupled to the ABM of surgical flow that reproduces the daily activity of a large surgical suite over a long period of time [30]. The model was calibrated using custom-made sensor systems placed at key locations of the surgical suite to capture the daily activity over a period of a year [28]. This OR suite, dedicated to general surgery, has about 20 ORs distributed in a layout as seen in Figure 13 and is rather typical of the activity in a large urban hospital. A simple stochastic model is assumed for the source of surgical smoke in each OR, similar to the previous one. The probability *p_door_* ∈ (0,1) of the number of OR door openings per minute is a parameter of the model. On average, one door opening every two minutes during a surgery is rather standard. This is mainly due to the fact that the staff may have to support logistics in various ORs at the same time, and that coordination of team activity is still done by a direct conversation in the surgical workflow. In fact, it is common knowledge that a door opening every 8 minutes on average would correspond to a very strict policy controlling traffic in the surgical suite, but it would only reduce the exposure in the hallway by half. From the simulation, it is concluded that long-term exposure to surgical smoke in the hallway is about the same order of magnitude as the one in the OR.

Figures 9 to 10 demonstrate the effect of the frequency of door openings on the average concentration of pollutants that a staff member is exposed to during the day. There was a noticeable low concentration at the upstream end of the halls, which was confirmed by direct measurement with the particle counter. Enforcing strict control on door openings may reduce the level of transport and diffusion of hazardous airborne particles in the hallway by half. The model can be ran to test a fictitious situation as in Figure 11 where every other OR has an ideal practice and generates no surgical smoke at all. The usage of ideal exhaust ventilation devices during surgery in half of the ORs has a direct linear correlation with the rate of exposure in the hallway, and it seems to be the most efficient technique to reduce long-term exposure; it cuts down the staff’s exposure to surgical smoke in the hallway by half.

**Figure 9.**
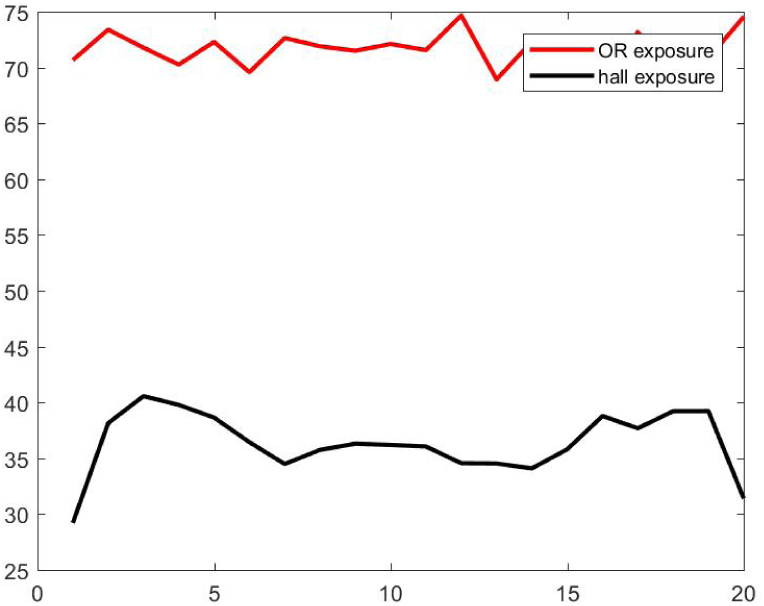
Effect of the frequency of door openings on the concentration of pollutants (y-axis) in each OR (x-axis) and in the respective portion of hallway. Here the frequency is set at 8 *min*.

**Figure 10.**
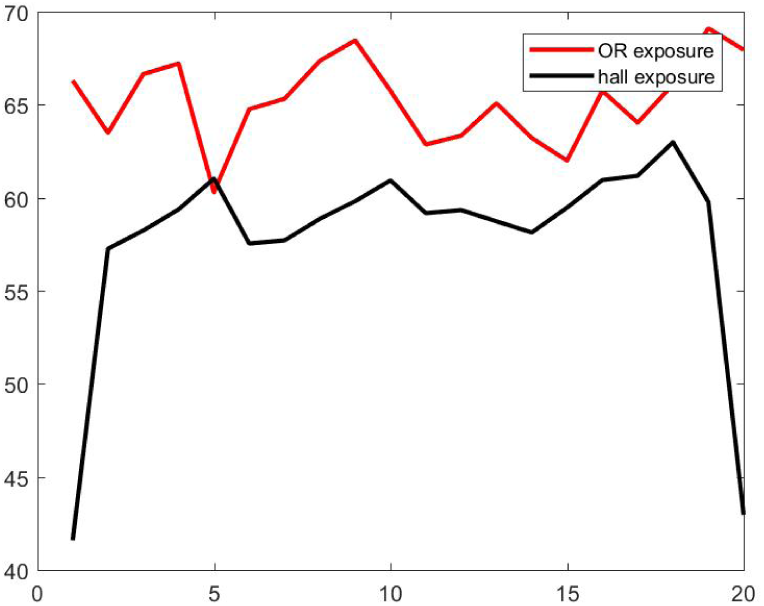
Effect of the frequency of door openings on the concentration of pollutants (y-axis) in each OR (x-axis) and in the respective portion of hallway. Here the frequency is set at 2 *min*.

**Figure 11.**
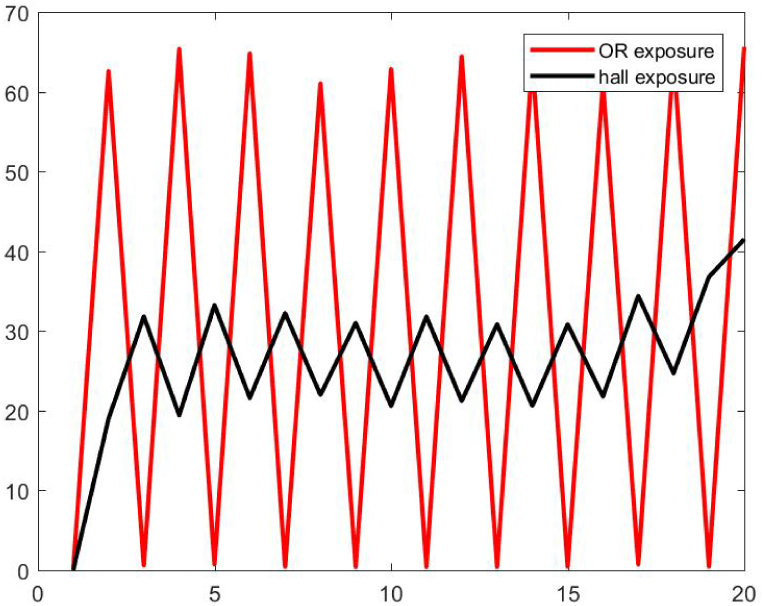
Hypothetical simulation where only half of the ORs produce surgical smoke. Here the frequency of door opening is set at 2 *min*.

## Discussion

About half a million healthcare professionals are exposed, daily, to surgical smoke in their clinical activities. Transport and diffusion of hazardous airborne particles such as virus, generated by surgical smoke in particular, and its long-term effects on staff have not been studied systematically yet.

The debate on the impact of surgical smoke on patients’ and staff’s health is reminiscent of the incident involving airborne hazards from anesthetic gas [22,33–36]. The national study led by the American Society of Anesthesiologists established that “female members in the operational room-exposed group were subject to increase risks of spontaneous absorption, congenital anomalies in their children, cancer and hepatic and renal disease.” While the link with Waste Anesthetic Gas (WAG) was not clearly established at that time, except in animal studies, the stream of work initiated in the 70’s [22,36] eventually ended up in a better management of WAG “by always using scavenging systems, by periodically testing anesthetic machines for gas leaks, and by not emptying or filling vaporizers” [37].

Unfortunately, the efficiency of surgical masks to prevent virus transmission or surgical smoke breath intake is usually tested using non-biological markers while their use in hospitals is mostly against airborne biological particles. Standard surgical masks and filtration techniques are not effective on UFP, which include viruses and bacteria. For example the SARS-CoV-2 as a size comprised between 0.06 to 0.14 microns [38]. Long *et al*. showed in a new meta-analysis that there was no significant difference in effectiveness between surgical masks and N95 masks against laboratory-confirmed respiratory viral infections [12] especially at higher inhalation flow rate [13]. Seongman *et al*. also showed that for viruses, effectiveness of surgical mask (and cotton based masks in the paper) are dependent to virus concentration and flow rate of inhalation but also showed a higher concentration of viral load on the outside of the mask compared to the inside [39]. Being able to know where these particles are and in what concentration seem then the best protection and awareness for staff to avoid staying too long in contact. Deposition rate of these particles is not addressed in this paper and researchers are still debating about lifetime on surface of the SARS-CoV-2 virus.

A method to construct a surgical-suite-specific model of the transport-diffusion of airborne particles can quickly be calibrated with cost-effective wireless particle counters. Coupling this indoor quality model to the previous multi-scale model of surgical flow [27–29] allows quantification of surgical smoke exposure across long periods of time and provides a rationale for recommendations. As a matter of fact, the ABM of surgical workflow allows insight into the human behavior factor, which can be included in the analysis. This work may expose rare events, such as contamination from one OR to another, which when accumulated over the months becomes a tangible risk. This study has potential because it can run for long periods of time and can address the complexity of hundreds of staff’s spatiotemporal behaviors in a large OR suite.

The CFD model requires detailed geometric and boundary conditions to be reliable; the *k* − *ϵ* model is an approximation that has its own limit as well. Running a CFD model is a tedious process both in setting up the mesh and simulation parameters, as well as in terms of central processing unit (CPU) time required. CFD was used here only to test the components of a hybrid stochastic compartment model that incorporates the mechanism of diffusion-transport of airborne particles at the surgical suite scale over a one-year period. A coarse statistical model was used for the source of surgical smoke: the actual generation of surgical smoke depends on the surgery team, type of procedure, and many more parameters. However, the capability to non-invasively monitor such parameters using appropriate sensors via the cyber-physical system is available. The hybrid Partial Differential Equation (PDE) compartment model provides a first-order approximation of average exposure at the room scale. The delay in the transmission conditions between the OR and the hall in Eq. 4 and 5 is not essential to reproduce the result on daily exposure to smoke. In the meantime, the uncertainty of the HVAC input/output provides a much larger error. Furthermore, deposition of particles on the OR’s surfaces was not taken into account. The deposition of UFP may be expected to be negligible. A low accuracy model that carries an error of the order of 20% could, however, be conclusive for this study.

It is particularly important to recognize the impact of door design and human behavior when considering hazardous airborne particles spreading throughout a surgical suite. OR doors constantly leak air depending on the difference of pressure with the outside hall, and they contribute to the transport of particles throughout the surgical suite. The door opening effect depends on the motion of the door and also the difference of temperature between the OR and the hallway. Some of these negative impacts can be controlled by a better design of the door and of the temperature control in order to work with a more cost-effective HVAC design. The benefit of positive pressure in the OR is still canceled by door openings, inducing possible back-flow and contamination from the hallway, especially when the door stays open for several minutes. Therefore, efficient movements by personnel may improve indoor air quality and should be quantified. The architectural design of the OR suite should optimize the circulation of staff and patient movement activities.

The next important step in the modeling to address the complementary aspect of biological transmission versus physical transportation is to correlate the database of staff’s pulmonary events with the study’s findings to recognize these rare events. We can then translate the quantitative model of surgical smoke transport into a risk assessment for staff’s health. The model should be surgery-specific: an efficient cyber-physical infrastructure should non-invasively monitor energy usage and smoke presence to instantly deliver awareness on practices that can improve air quality. A factor that has been neglected is the deposition of surgical smoke in the common storage area, see Figure 13, in which all ORs have personal access to via their back-doors. This may offer a different mechanism of propagation of biological material.

In the current study, quantification of surgical smoke concentration in the hallway, the duration of exposure along the year, and the mechanism of propagation of hazardous airborne particles from one OR to another was feasible. On the practical side, an automatic sliding OR door seems to be a better solution over a traditional door’s rotation that acts as a pump. The analysis can also be extended to address the problem of the optimum placement of UV lights in the hallway to improve air quality in an efficient and controlled way. Finally, the importance of AORN’s guideline in the use of a vacuum system during surgery needs to be reinforced at a time when elective surgery may involve asymptomatic COVID-19 patients.

## Methods

### Computational Air Flow Simulation of the Operating Room

To simulate the airflow and dispersion of surgical smoke, an OR that is representative of the surgical suite shown in Figure 13 was used. Measurements for calibrations and verifications were conducted in a real OR of this dimension when there were no surgeries taking place. Figure 1 provides the schematic of all boundary conditions and geometric parameters. This CFD approach is used to justify and build a simplified large-scale model of the airflow in the surgical suite. A 3D Cartesian coordinate system was used with length along the *x*-direction, width along the *y*-direction, and height along the *z*-direction. The OR is 7.5 *m* long, 6 *m* wide, and has a height of 2.7 *m* (see Figure 1). The model takes into account the architecture of the room, the operating table location, and the HVAC system design in the OR as well as in the hallway. The corridor was modeled as a rectangle of 12 *m* long, 2.5 *m* wide and a height of 2.7 *m*. The operating table is displayed as a rectangle in the middle of the OR, and the anesthesia equipment is also simulated by a rectangle that is close to the table. The computation of the flow was done by using a pressure based solver and an ANSYS Fluent solver in steady states first and then transient mode after. The model’s geometry was meshed using an unstructured tetrahedral grid with about 10^6^ elements. The exact size of the mesh depends on the angle of the door with its initial closed position since the mesh gets refined at the interfaces. The airflow is assumed to be turbulent [24] and was modeled using the *k* − *ϵ* turbulence model, taking into account gravity to introduce the Boussinesq approximation in the Navier-Stokes equation. It is the most common model for indoor airflow simulation in which the turbulent kinetic energy *k* and turbulent dissipation rate *ϵ* are modeled. The temperature and pressure boundary conditions in the model were measured, as reported in Table 2. Typically, the OR is kept cooler than the hallway, and the inlet vent inside the OR blows air at a temperature as low as 13°C.

To match the ventilation infrastructure, the model has three different inlet vents in the ceiling. In principle, there are two rows of vents on the left and right of the middle vents that are present to avoid any flow returning towards the operating table. The slightly different velocity flows for each inlet were provided as stated in Table 2 in order to replicate the anemometry measurement (via Peak Meter MS6252B) obtained near these inlets – it was noticed that the large surgical lights over the surgery field have the tendency to obstruct part of the inlet flow. The OR also contains two outlet vents that suck out the air in the OR with such a velocity that the outlets pump out less volume than the volume injected by the ceiling inlets, which creates a positive pressure of about 8 Pa. Pressurization is a key factor in controlling room airflow patterns in a healthcare facility. Positive pressurization is used to maintain airflow from clean to less-clean spaces. The appropriate airflow offset to reach the desired pressure differential depends mostly on the quality of the construction of the room. It is difficult, if not impossible, to know what the room’s leakage area is before finishing the construction and doing measurements of airflow. The Facility Management Service (FMS) of the hospital was able to supply the values of volume per minute that the inlets are blowing (1.15 *m*^3^*/s*) and of the volume going through the outlets (0.55 *m*^3^*/s*). As the surface of the outlets is known, the velocity of the outlet vents were 0.5 *m/s* for the left one and 0.4 *m/s* for the right one, reported on Figure 1. The volume of extra air in the OR is the difference: 1.15 − 0.55 = 0.6 *m*^3^*/s*.

Due to the positive pressure of 8 Pa present in the OR, this additional volume leaks out of the OR through either the door being left open or the narrow gap around the door when it is closed. In that case, the free boundary surface was estimated to be 8.5 *cm*^2^.

For the hallway, a uniform inflow boundary condition of 0.1 *m/s* was imposed in order to take into account the anemometry measurement mentioned above. This upstream boundary condition is completed by a free outlet boundary condition at the other end of the hallway. To be as realistic as possible, the two existing inlet vents’ boundary conditions were respectively added for the inlet vent located on the ceiling and the other inlet located close to the entry door of the next OR, both with a velocity of 1.2 *m/s*. The goal of the CFD simulation is to be able to assess the rate of pollutant leaving or entering the OR depending on if the door is closed (0°) or completely open (90°). During the observation of the clinic work, OR doors would sometimes stay open for several minutes and for different reasons. The doors of the ORs downstream were left open in order to check the potential inflow of pollutants from the other ORs.

To validate the model, measurements were done of velocity flow and of the concentration of particles at various locations that were close to specific regions of interest – the doorframe location in particular (see Table 3). It is unrealistic, in practice, to build a CFD model of the whole surgical suite and run this model for an extensive period of time. Next, an upscale model will be presented that will use the present CFD simulation to verify some of the key parameter values, especially relating to transmission parameters between ORs and the hallway.

### Hybrid Box-PDE Model of Airborne Contamination in the Surgical Suite

As an example, consider a system of 10 identical ORs aligned on one side of a hallway. Each OR has one door access to the hallway. This system is part of a standard OR suite and represents one half of the facility in Figure 13 that has an architectural design almost symmetric with its two circulation side-halls. Computing the concentration of a so-called “marker,” which can be a specific gas or set of airborne particles in the air of this OR suite, is particularly of interest. This marker is generated from the location of the OR’s surgical table, where a surgeon is using an electrosurgical instrument that produces smoke from the thermal destruction of tissues. The marker can also be the particles resulting from evaporation of any alcohol-based chemical used either to prep the patient or to clean the OR. The model has two parts: first, a compartment-like model that can monitor the indoor pollution [40]; second, a multi-scale ABM that simulates the surgical flow activity and the impact on the indoor air quality either from the source of surgical smoke or from door openings affecting the dispersion of pollutants [30]. Staff movement throughout the OR suite via door openings and closings will manifestly be a key mechanism for propagation of markers. The indoor air quality is a linear set of differential equations that will be slightly more complex than a standard compartment model since the coefficient will be stochastic, the sources and output/leaks of the particles term will have a time delay built in, and the hallway will require a transport equation.

The rationale for building this specific model will come out of the set of experiments described hereafter. Next, the description of the acquisition process to identify the production of airborne contaminants will be explained.

### Experimental Setup to Assess the Source of Particles and Smoke

For this experiment in a surgical training facility, electrosurgical energy was delivered on the surface of two pieces of pork meat, each 2 *cm* thick, placed on an OR table. Three types of energy delivery systems were compared: electrosurgery (conduction) via the Covidien ForceTriad monopolar device, ultrasonic (mechanic) with the Ethicon Harmonic Scalpel P06674 device, and laser tissue ablation with Erbe APC (Argon Plasma Coagulation) 2 device. To keep the tissue burn superficial, a pattern of parallel lines was followed with each device and always used unburned areas of the meat. The energy was delivered for a period of 30s up to 60s in order to produce a large quantity of smoke and thus particles. The measurement was done by several laser particle counters from Dylos Corp placed at various distances from the source (http://www.dylosproducts.com/dc1700.html). They give an average particle count every minute in a unit system with units *u_d_* that correspond to 0.01 particles per cubic foot. A traditional problem with the validation of particle count in laboratory conditions is that particles are not all the same uniform size. According to smartAir (http://smartairfilters.com/cn/en/), the Dylos system output is highly correlated (r = 0.8) to a “ground true” measurement provided by a high-end system such as the Sibata LD 6S that is claimed to be accurate within 10% in controlled laboratory conditions. According to smartAir, the Dylos system seems particularly accurate at the lower concentration ends, which is of interest for this study’s purpose. Semple *et al*. [41] also compared the Dylos system with a more expensive system: the Sidepak AM510 Personal Aerosol Monitors (TSI, Minnesota, USA). They concluded that the Dylos’ output agrees closely with the one produced by the Sidepak instrument with a mean difference of 0.09 *μg/m*^3^.

The Dylos sensors were set up to track particles of small size in the range from 0.5 to 2.5 *microns*, which are the sizes of biological material. The results were checked systematically by comparing the measures of several sensors at the same location to show consistency, as well as checked that the particle count lowers back down to nearly zero in a clean-air room with AC equipped with High Efficiency Particulate Air (HEPA) filters. Each experiment was started from an initial clean-air condition of a small particle count, fewer than 50 units, which is much less than the number of particles counted during energy delivery. It took about 6 minutes to reach the initial clear-air count after each experiment.

For each experiment, the concentration increased to a maximum after a short time delay *s* from the time the energy was delivered; this delay depends on the distance to the source. The concentration then exponentially relaxes to zero in time. Consequently, the model of source dispersion is an exponential function as follows:

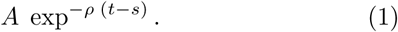

The least squares fitting technique was used to interpolate the data with this function. The amplitude of the source A (see Fig. 12), the delay *s* on particle diffusion and transport to reach the sensor and the rate of “diffusion decay” *ρ >* 0 were identified. The accuracy on *s*, which measures the time interval between the source production and the peak of the signal, cannot be faster than one minute since the sensor only works at a one-minute timescale. A delay of *s* ≤ 1 was found to be a good approximation for all three energy devices. Each experiment was done 4 to 5 times depending on the variability of the results. Therefore, about 24 to 30 data points were available to identify the parameters *A*, *s*, and *ρ* for each energy device that was tested.

**Figure 12.**
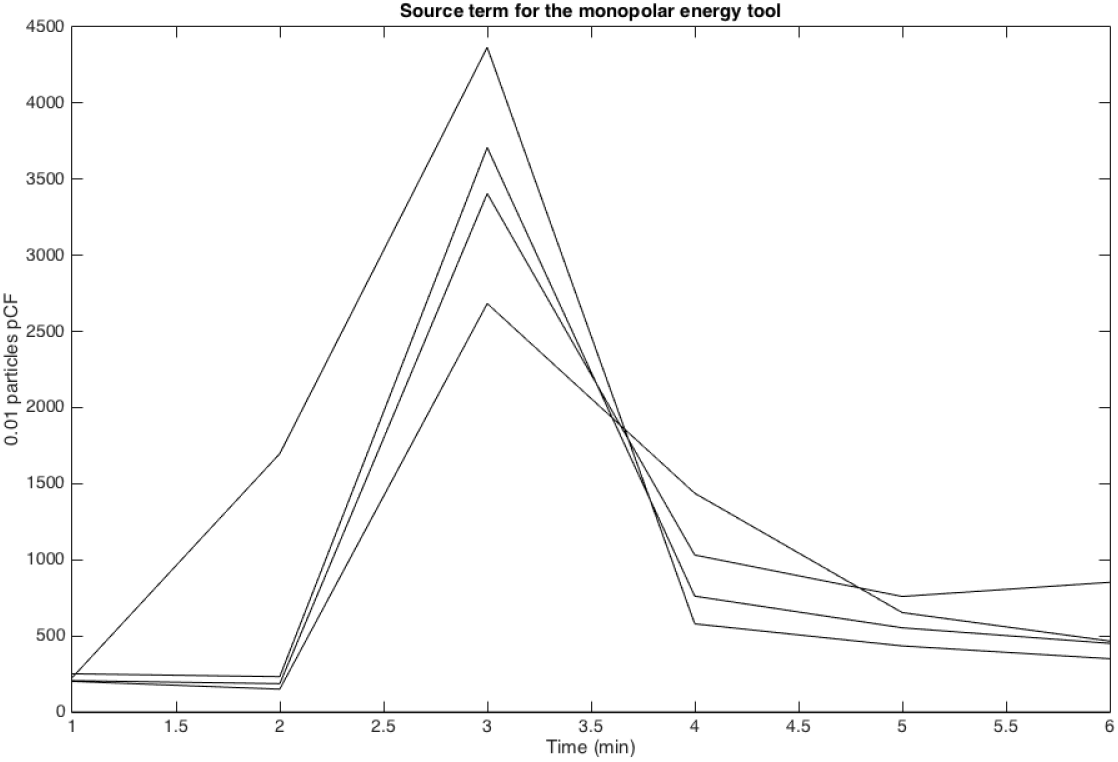
Dynamic of particle count with the monopolar energy system

Now, the protocol experiment to assess the transport and diffusion of particles in different areas of a large OR suite will be described.

### Assimilation of Experimental Data

This set of experiments, as opposed to the previous one, was done in a large OR suite late at night and on weekends when the ORs were empty and had clean air with high-efficiency HVAC. A hairspray product (Lamaur Vitae, unscented) was used as the marker and sprayed for a duration of 1 to 2 seconds to track its small particles while keeping the same positions of the Dylos systems. The experiment first tested the propagation in a closed-door OR with the source above the OR surgical table. The spray nozzle was held facing the near-vertical direction, pointing to the ceiling. A distribution of sensors as displayed in Figure 3 was used.

The initial observation was that all four sensors distributed along the central line of the whole OR space were getting a particle count of the same order of magnitude after an average of 15 seconds. The mixing of particles was quite extensive within a minute by reason of the HVAC input/output design in the OR, and that the concentrations on each sensor quickly relaxed to zero. This observation is also coherent with the results of the CFD model of the flow circulation described above.

A method identical to the previous one was used to identify the key parameters *A, s*, and *ρ* characteristic of the dispersion of hairspray in the OR.

The model for OR diffusion of particles is then

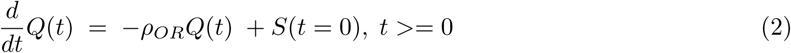

where S(t = 0) denotes the source production that is non zero at time zero. This simple Ordinary Differential Equation (ODE) model provides an average of particle concentration in the OR at the minute timescale.

A first-order implicit Euler scheme with a time step *dt* of one minute is used:

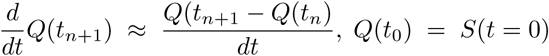

An entirely similar technique is used to describe the dynamic of particle diffusion and transport in the hallway, except that the hallway is discretized as a one-dimensional structure of consecutive hall blocks located at the same level as the OR block. In this part of the experiment, the source is set in the hallway – see Figure 13.

**Figure 13.**
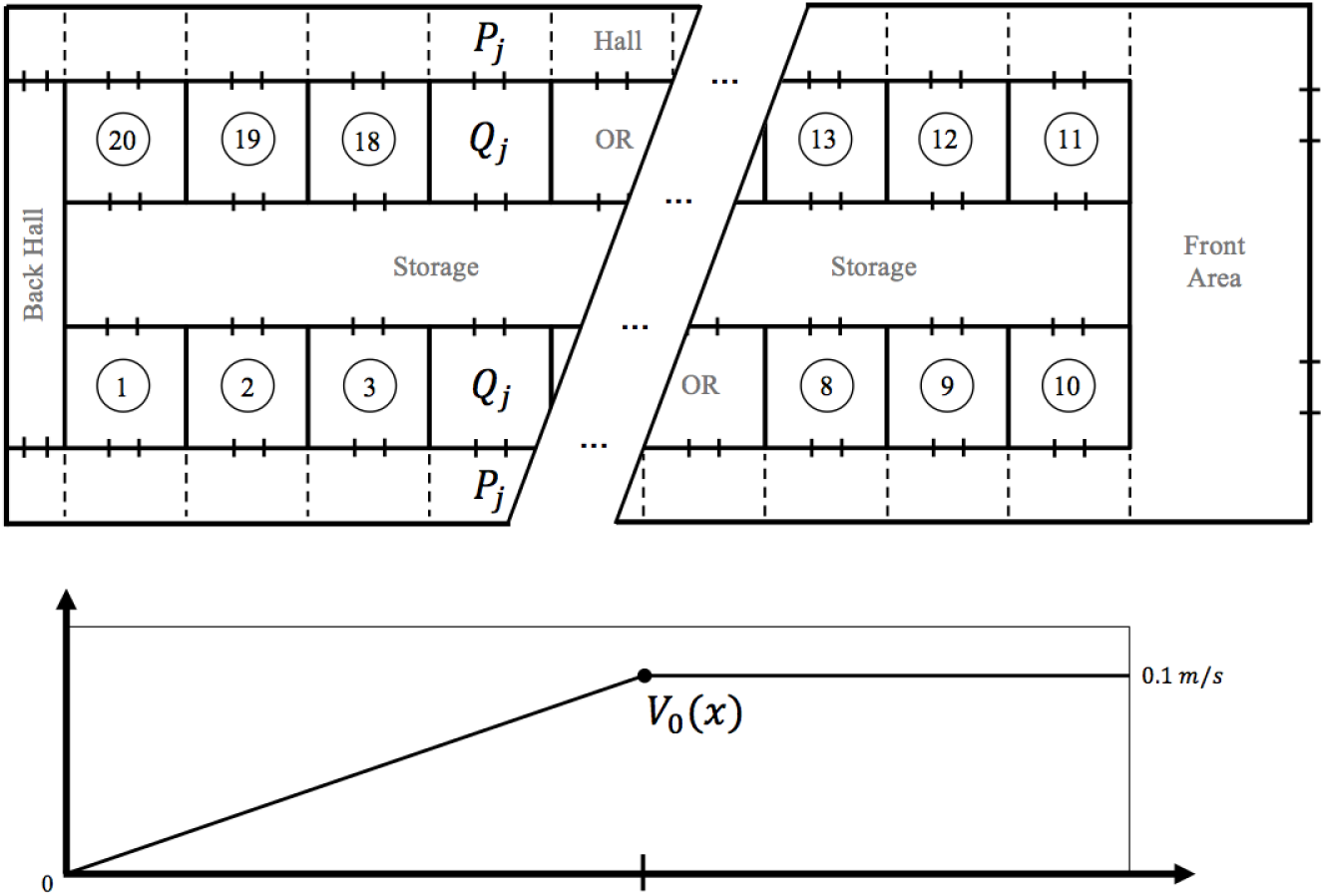
Approximate floor plan of the surgical suite used during the clinical test. The dash lines represent the coarse finite volume grid used for the diffusion transport equation in the hallway. *v_0_*(*x*) is the velocity of the airflow moving down the hallway.

As noticed earlier, there is a slow but significant air flow speed v_0_ in the hall, pointing in the direction of the main entrance of the surgical suite, situated on the right of the map in Figure 13. Naturally, the high pressure of the OR is designed to drive the airflow out and the front corridor seems to be a significant outlet. On the opposite end of the hall, situated on the left of the map, Figure 13, this velocity is close to zero. It is assumed that *v*_0_(*x*) is an affine function, with a linear growth from 0 to 0.1*m*/*s* at mid-hall, and a constant value beyond. The model of hallway diffusion of particles is then

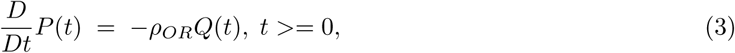

where 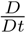 denotes the total derivative 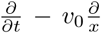 using the *x* coordinate system in the one space dimension hall model.

To assess the transmission of particles from an OR to the adjacent hallway with closed OR doors, the same experiments were run with some of the sensors placed in the hallway either facing the closed door or sitting at a location in the hallway (see Figure 3). As a matter of fact, the door of the OR is not perfectly sealed due to the difference between the pressure inside the OR and the lower pressure in the hallway, a significant airflow with velocity around the order of 1 *m*/*s* exists at the gap located between the door’s edge and the door frame. Table 3 provides the values obtained both from measurement and the CFD simulation. A similar technique is used to represent the diffusion coefficient as well as the delay s that is now interpreted as the time it takes for the particles to flow from the OR to the hallway right outside the door. This transmission condition will be entered into the model to couple equation 2 and 3.

Finally, an entirely similar approach is used to get the transmission in the compartment model when the door of a specific OR is wide open. In such case, the gradient of pressure between the OR and the hallway nearly vanishes, and only the diffusion process is dominant in that section of the hallway.

Now, the simple compartment-like model to monitor, in time and in space, the diffusion and transport of particles with intermittent source production in each OR will be assembled. Such a source of pollutants corresponds to either the use of some chemicals or the use of electrosurgical instruments during surgery. The goal is to get the average rate at which the staff working in the OR suite is getting exposed to particle concentration emanating from surgical smoke throughout the day. Potential propagation of particles that may carry biological material from one OR to another is also of interest.

### Surgical Suite Model as a System

As discussed earlier, the concentration is tracked in time and in space with a coarse time step of one minute. This time step scale is coherent with the measurement system used for particle counting. One minute is also roughly the time that the particles emitted from a point source next to the OR table need to transport and diffuse throughout the OR block once released. The compartment model computes the global concentration of the particles in each OR as well as in each section of the hall adjacent to the OR. These concentrations are denoted respectively *Q_j_* (*t*) for OR number *j* at time *t* and *P_j_*(*t*) for the corresponding section of the hall – see Figure 13.

The source of particles is denoted as *S_j_* (*t*). In principle, *S_j_* (*t*) should be non-zero for a limited period of time and follow a statistical model based on the different phases of the surgery and the knowledge of electrosurgical instrument used during a surgical procedure [18]. The coefficients of decay are defined inside different parts of the model (*ρ_OR_* and *ρ_Hall_*) as well as the coefficients of transmission between these spaces (*α_OR_* from the OR to the hall and *γ_Hall_* for the opposite). *β_OR_* represents the flow from the OR to the hall when the door is open. The frequency of door openings is following a statistical model based on where the surgery is at; 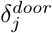 is a function of time and is 1 if the door is open, 0 otherwise. The simulation of the surgery schedule uses data from the SmartOR project [42], which will be detailed later on. Only the door openings of the order of a minute will be counted and *γ_Hall_* = *β_OR_* will be assumed because the gradient of pressure between the OR and hallway vanishes.

The system model of marker transport-diffusion in the OR suite is:

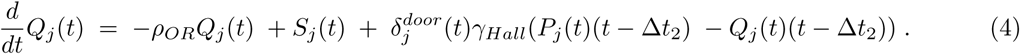

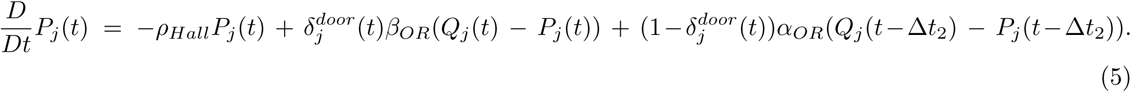

An additional unknown to track back-flow of marker in the OR coming from the hallway can be introduced with:

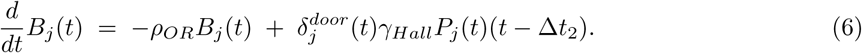

Using this equation, the number of particles going from one OR to another can be separately counted. This number is expected to be very low – see “Results” section.

The model (4, 5) is not a standard box model. First, the source term has a delay built-in to simulate the transmission conditions observed. Second, equation (5) is a PDE, more precisely a linear transport equation. Third, most of the coefficients are stochastic, especially those related to door openings and sources that are linked to human behavior. Because the system of equations is linear, the superposition principle has been implicitly used to retrieve each unknown coefficient from the experimental protocol.

Let us describe our surgical flow model more precisely in order to provide an accurate description on how we manage to compute the source term *S_j_* (*t*).

For each of the standard OR stages of the surgery, an attributed *State* value is given as follows:

- Phase 1: anesthesia preparation label as *State=1*.
- Phase 2: surgical preparation to access as *State=2*.
- Phase 3: surgery procedure as *State=3*.
- Phase 4: surgery closing as *State=2*.
- Phase 5: ending anesthesia as *State=1*.
- Phase 6: room in the process of cleaning or free as *State=0*.

The type of airborne marker expected to release depends on those *State* values. For example: In *State 0*, cleaning crew uses a lot of chemical products that quickly evaporate in the OR. Similarly, a different type of sterilization product is used to prep the patient in *State 1*. In *State 2*, cauterization is often used for a short period of time. In *State 3*, various phases of the surgery will require energy delivery instruments to cut tissue and access specific anatomy or tumors.

A stochastic model of energy delivery is used that consists of delivering short time fractions of energy in several consecutive minutes. The parameters of that model are: the frequency of energy delivery denoted *f*, the duration of the impulse denoted ξ, and the number of repetition *r*. A uniform probabilistic distribution of events is used within these intervals of variation for each parameter.

Figure 14 provides a typical example of the number of door openings observed in the OR at a 15-minute interval. Both the detection of door openings and a patient bed coming in and out were provided by the sensors of the cyber-physical infrastructure [27]. A stochastic model of door openings will be used based on a uniform frequency of door opening during surgery, even though this distribution is non-uniform in practice and tends to concentrate at the beginning and the end of a case.

**Figure 14.**
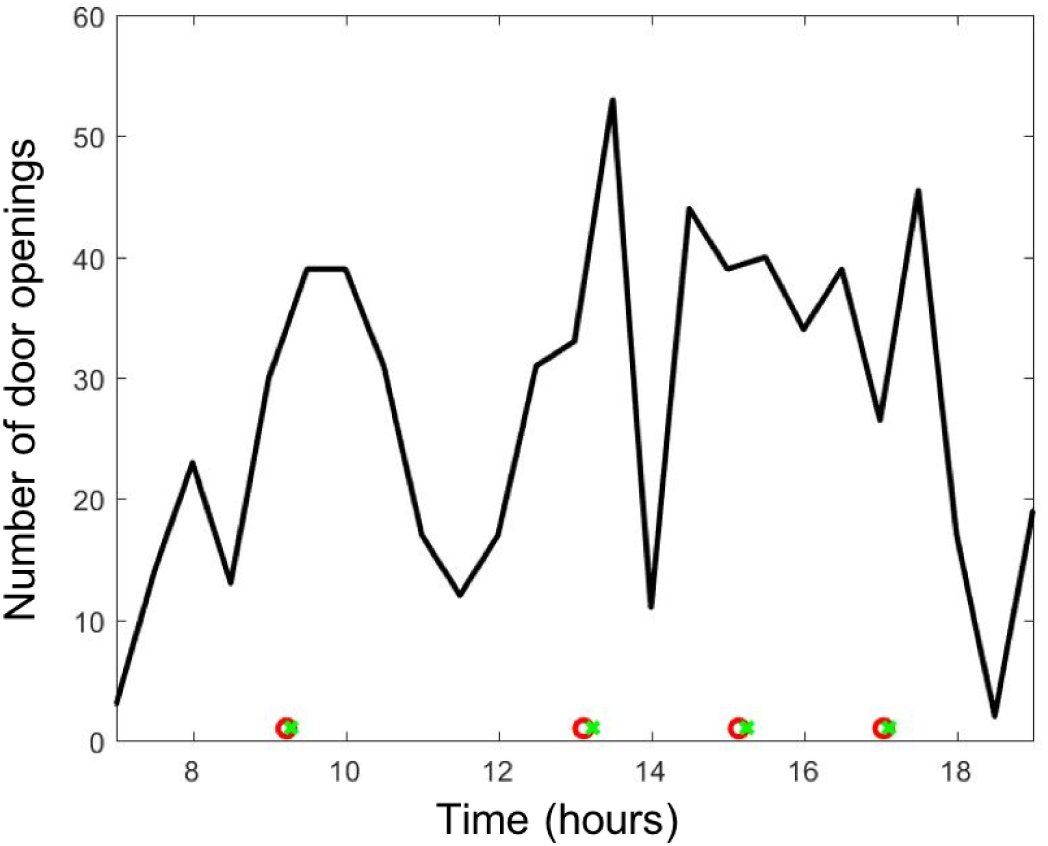
Plot of the number of door openings every 15 minutes during a clinical day from 7 a.m. to 7 p.m.; ○ and x on the horizontal axis corresponds respectively to the entering and exiting time of the bed of the patient. This example has two procedures.

The model of air pollution in the surgical suite will first be tested with a simplified model of surgical flow as follows: to provide the timeline of events, the model assumes there are three surgical procedures in each OR. The timeline of each surgery will be such that: Phase 1 and Phase 5 last 12.5*min* ± 5*min*, Phase 2 and Phase 4 last 15*min* ± 5*min*, Phase 3 is the surgery itself that lasts 65*min ±* 25*min*. Phase 6 corresponds to a turnover time between surgeries that lasts 30*min ±* 10*min*. This simplified model of surgery scheduling has the correct order of time-length for each phase. Its simplicity allows a sensitivity analysis to run with respect to the key parameters of the indoor air quality model that can be easily interpreted.

Next, the model will be coupled on an ABM of an existing large general-surgery suite in the hospital that has been calibrated by tracking about 1000 procedures over one-year [30]. This model is complex and specific to a 20 ORs surgical suite of a 1700 bed hospital, which has been monitored for over two years.

### Clinical Model of Airborne Hazard and Integration of Human Behavior in the Simulation

To asses, the impact of human behavior on the transport and diffusion of surgical smoke in the surgical suite over a period of one year, a realistic ABM of the surgical flow and of people behavior is now used. A byproduct of this study is the assessment of the air quality and the risk factors associated with surgical smoke by coupling it to the present model of transport and diffusion of airborne particles generated by surgical smoke.

The method to construct this model is briefly explained in this paragraph. The exact description of the model goes beyond the scope of this paper’s focus on air quality and has been detailed in Garbey et al. [30]. The mathematical model of surgical flow is built upon observations and robust clinical data covering 1000 procedures with a noninvasive array of sensors that automatically monitor the surgical flow. To this end, several ORs were equipped with sensors that capture timestamps [27, 28,43] corresponding to the different states described in the previous section.

Overall, the model can simulate the OR status of a large surgical suite during any clinical day and can be run over a long period of time. The model is able to reproduce the statistic distribution pattern over a year of performance indicators: turnover time, induction of anesthesia time, the time between extubation, and patient exit. The model classifies the human factors’ impact and limitation of shared resources on flow efficiency. In the end, communication delays and sub-optimal OR awareness in large surgical suites have significant impacts on performance and should be addressed. This paper concentrates on the duration of surgery *State* 3 that corresponds to how long surgical smoke is generated, and how behavior inducing OR door openings are responsible in part for the spread of surgical smoke and other agents. The output of the ABM model of surgical flow coupled to the air quality model is the number of hours per year that staff gets exposed to surgical smoke in the OR and hallway. Various scenarios have been run related to the rate of adoption of vacuum systems for surgical smoke and OR door openings to discuss the influence of human behavior on those results.

## Data Availability

All data generated and analysed during this study are included in this published article. Additional information can be available from the corresponding author on reasonable request.

## Author contributions statement

This project is highly interdisciplinary and required all co-authors contributions to establish the concept and reach the goal of the paper.

M.G. leads the project, designed the overall framework, including the hybrid model, agent-based clinical model, and Matlab code implementation.

G.J. did the CFD model at the OR scale, participated in the design of the overall method and ran the experiments with air quality sensors required for validation.

S.F. participated in the experiments with air quality sensors and with OR door activity sensors.

All authors contributed to the redaction of this publication.

## Additional information

The authors declare no competing interests. All data generated and analysed during this study are included in this published article. Additional information can be available from the corresponding author on reasonable request.

## Notes

### Competing Interest Statement

The authors have declared no competing interest.

### Funding Statement

No external funding was received.

## References

1. Morawska L, Cao J. Airborne transmission of SARS-CoV-2: The world should face the reality. Environment International. 2020; p. 105730.

2. Bourouiba L. Turbulent gas clouds and respiratory pathogen emissions: potential implications for reducing transmission of COVID-19. Jama. 2020;.

3. Liu Y, Song Y, Hu X, Yan L, Zhu X. Review Awareness of surgical smoke hazards and enhancement of surgical smoke prevention among the gynecologists. Journal of Cancer. 2019;10(12):2788–2799.

4. Barrett WL, Garber SM. Surgical smoke—a review of the literature: Is this just a lot of hot air? Surg Endosc. 2003;17(x):979–987.

5. Hill D, O’Neill J, Powell R, Oliver D. Surgical smoke – a health hazard in the operating theatre: a study to quantify exposure and a survey of the use of smoke extractor systems in UK plastic surgery units. J Plast Reconstr Aesthet Surg. 2012;65(7):911–916.

6. Steege A, Boiano J, Sweeney M. Secondhand smoke in the operating room? Precautionary practices lacking for surgical smoke. Am J Ind Med. 2016;59(11):1020–1031.

7. Nezhat C, Winer W, Nezhat F, et al. Smoke from Laser Surgery: Is there a health hazard? Lasers in Surgery and Medicine. 1987;7(x):376–382.

8. Garden J, O’Banion K, Shelnitz L, et al. Papillomavirus in the vapor of carbon dioxide laser – treated verrucae. Journal of American Medical Association. 1988;259(x):1199–1202.

9. Romano F, Gusten J, De Antonellis S, Joppolo CM. Electrosurgical Smoke: Ultrafine Particle Measurements and Work Environment Quality in Different Operating Theatres. Int J Environ Res Public Health. 2017;14(137):doi:10.3390/ijerph14020137.

10. Sisler JD, Shaffer J, Soo JC, LeBouf RFL, Harper M, Qian Y, et al. In vitro toxicological evaluation of surgical smoke from human tissue. Journal of Occupational Medicine and Toxicology. 2018;13(12):https://doi.org/10.1186/s12995-018-0193-x.

11. Wu X, Nethery RC, Sabath BM, Braun D, Dominici F. Exposure to air pollution and COVID-19 mortality in the United States: A nationwide cross-sectional study. medRxiv. 2020;doi:10.1101/2020.04.05.20054502.

12. Long Y, Hu T, Liu L, Chen R, Guo Q, Yang L, et al. Effectiveness of N95 respirators versus surgical masks against influenza: A systematic review and meta-analysis. Journal of Evidence-Based Medicine. 2020;.

13. Bałazy A, Toivola M, Adhikari A, Sivasubramani SK, Reponen T, Grinshpun SA. Do N95 respirators provide 95% protection level against airborne viruses, and how adequate are surgical masks? American journal of infection control. 2006;34(2):51–57.

14. Stocks GW, Self SD, Thompson B, Adame XA, O’Connor DP. Predicting bacterial populations based on airborne particulates: A study performed in nonlaminar flow operating rooms during joint arthroplasty surgery. American Journal of Infection Control. 2010;38(3):199—204.

15. Cristina ML, Spagnolo AM, Sartini M, Panatto D, Gasparini R, Orlando P, et al. Can particulate air sampling predict microbial load in operating theatres for arthroplasty? PLoS One. 2012;7(12):e52809.

16. Lu J, Gu J, Li K, Xu C, Su W, Lai Z, et al. COVID-19 Outbreak Associated with Air Conditioning in Restaurant, Guangzhou, China, 2020. Emerging Infectious Diseases. 2020;26(7).

17. Ong SWX, Tan YK, Chia PY, Lee TH, Ng OT, Wong MSY, et al. Air, surface environmental, and personal protective equipment contamination by severe acute respiratory syndrome coronavirus 2 (SARS-CoV-2) from a symptomatic patient. Jama. 2020;.

18. Meeuwsen F, Guédon A, Arkenbout E, van der Elst M, Dankelman J, van den Dobbelsteen J. The Art of Electrosurgery: Trainees and Experts. Surg Innov. 2017;24(4):373–378.

19. Tang J, Nicolle A, Pantelic J, Klettner C, Su R, Kalliomaki P, et al. Different types of door-opening motions as contributing factors to containment failures in hospital isolation rooms. PLoS One. 2013;8(6):e66663.

20. Eames I, Shoaib D, Klettner CA, Taban V. Movement of airborne contaminants in a hospital isolation room. J R Soc Interface. 2009;6(x):757–766.

21. Mousavi ES, Grosskopf KR. Airflow patterns due to door motion and pressurization in hospital isolation rooms. Science and Technology for the Built Environment Volume. 2016;22(4):379–384.

22. Cohen E, Bellville J, Brown BJ. Anesthesia, pregnancy, and miscarriage: a study of operating room nurses and anesthetists. Anesthesiology. 1971;35(4):343–7.

23. Emmerich S, Heinzerling D, Choi Ji, Persily A. Multizone Modeling of Strategies to Reduce the Spread of Airborne Infectious Agents in Healthcare Facilities. Building and Environment. 2013;60(x):105–115.

24. Awbi HB. Ventilation of buildingsy, 2nd ed. London; Spon Press; 2003.

25. Romano F, Aroccoa L, Gustenb J, Joppoloa CM. Numerical and experimental analysis of airborne particles control in an operating theater. Building and Environment. 2015;89(x):369–379.

26. Cincinelli A, Martellini T. Editorial: Indoor Air Quality and Health. Int J Environ Res Public Health. 2017;14(x):1286.

27. Joerger G, Rambourg J, Gaspard-Boulinc H, Conversy S, Bass BL, Dunkin BJ, et al. A CyberPhysical System to Improve the Management of a Large Suite of Operating Rooms. ACM Transactions on Cyber-Physical Systems. 2018;2(4):34.

28. Huang AY, Joerger G, Fikfak V, Salmon R, Dunkin BJ, Bass BL, et al. The SmartOR: a distributed sensor network to improve operating room efficiency. Surgical endoscopy. 2017;31(9):3590–3595.

29. Huang AY, Joerger G, Salmon R, Dunkin B, Sherman V, Bass BL, et al. A robust and non-obtrusive automatic event tracking system for operating room management to improve patient care. Surgical endoscopy. 2016;30(x):3638–3645.

30. Garbey M, Joerger G, Rambourg J, Dunkin B, Bass B. Multiscale Modeling of Surgical Flow in a Large Operating Room Suite: Understanding the Mechanism of Accumulation of Delays in Clinical Practice. Procedia Computer Science. 2017;108(x):1863–1872.

31. Weld KJ, Dryer S, Ames CD, Cho K, Hogan C, Lee M, et al. Analysis of Surgical Smoke Produced by Various Energy-Based Instruments and Effect on Laparoscopic Visibility. Journal of Endourology. 2007;21(3):347–351.

32. Flaniken B. Engineer’s HVAC Handbook. Proce; 2011.

33. Ad Hoc Committee on the effect of trace anesthetics on health of operating room personnel ASoA. Occupational disease among operating room personnel: national study. Anesthesiology. 1974;41(4):321–40.

34. Spence A, Cohen E, Brown BJ, Knill-Jones R, Himmelberger D. Occupational hazards for operating room–based physicians. Analysis of data from the United States and the United Kingdom. JAMA. 1977;238(x):955–9.

35. Hemminki K, Kyyrönen P, Lindbohm M. Spontaneous abortions and malformations in the offspring of nurses exposed to anaesthetic gases, cytostatic drugs, and other potential hazards in hospitals, based on registered information of outcome. J Epidemiol Community Health. 1985;39(2):141–7.

36. Rosenberg P, Kirves A. Miscarriages among operating theatre staff. Acta Anaesthesiol Scand Suppl. 1973;53(x):37–42.

37. Shuhaiber S, Koren G. Occupational exposure to inhaled anesthetic Is it a concern for pregnant women? Canadian Family Physician. 2000;46(x):2391–2392.

38. Zhu N, Zhang D, Wang W, Li X, Yang B, Song J, et al. A novel coronavirus from patients with pneumonia in China, 2019. New England Journal of Medicine. 2020;.

39. Bae S, Kim MC, Kim JY, Cha HH, Lim JS, Jung J, et al. Effectiveness of Surgical and Cotton Masks in Blocking SARS–CoV-2: A Controlled Comparison in 4 Patients. Annals of Internal Medicine. 2020;doi:10.7326/M20-1342.

40. research Council N, et al. Indoor pollutants. National Academies; 1981.

41. Semple S, Ibrahim A, Apsley A, Steiner M, Turner S. Using a new, low-cost air quality sensor to quantify second-hand smoke (SHS) levels in homes. Tob Control. 2015;2(x):153–8.

42. Huang AY, Joerger G, Salmon R, Dunkin B, Sherman V, Bass BL, et al. A robust and non-obtrusive automatic event tracking system for operating room management to improve patient care. Surgical endoscopy. 2015; p. 1–8.

43. Garbey M, Joerger G, Huang A, Salmon R, Kim J, Sherman V, et al. An intelligent hospital operating room to improve patient health care. Journal of Computational Surgery. 2015;2(1):1–10.

